# Facial and vocal markers of schizophrenia measured using remote smartphone assessments

**DOI:** 10.1101/2020.12.02.20219741

**Authors:** Isaac R. Galatzer-Levy, Anzar Abbas, Vidya Koesmahargyo, Vijay Yadav, M. Mercedes Perez-Rodriguez, Paul Rosenfield, Omkar Patil, Marissa F. Dockendorf, Matthew Moyer, Lisa A. Shipley, Bryan J. Hansen

**Affiliations:** AiCure, New York, NY; Psychiatry, New York University School of Medicine, New York, NY; Psychiatry, Icahn School of Medicine at Mount Sinai, New York, NY; Merck & Co., Inc., Kenilworth, NJ, USA

**Keywords:** digital biomarkers, phenotyping, computer-vision, facial expressivity, negative symptoms, vocal acoustics

## Abstract

**Background:** Machine learning-based facial and vocal measurements have demonstrated relationships with schizophrenia diagnosis and severity. Here, we determine their accuracy of when acquired through automated assessments conducted remotely through smartphones. Demonstrating utility and validity of remote and automated assessments conducted outside of controlled experimental settings can facilitate scaling such measurement tools to aid in risk assessment and tracking of treatment response in difficult to engage populations.

**Methods:** Measurements of facial and vocal characteristics including facial expressivity, vocal acoustics, and speech prevalence were assessed in 20 schizophrenia patients over the course of 2 weeks in response to two classes of prompts previously utilized in experimental laboratory assessments: *evoked* prompts, where subjects are guided to produce specific facial expressions and phonations, and *spontaneous* prompts, where subjects are presented stimuli in the form of emotionally evocative imagery and asked to freely respond. Facial and vocal measurements were assessed in relation to schizophrenia symptom severity using the Positive and Negative Syndrome Scale.

**Results:** Vocal markers including speech prevalence, vocal jitter, fundamental frequency, and vocal intensity demonstrated specificity as markers of negative symptom severity while measurement of facial expressivity demonstrated itself as a robust marker of overall schizophrenia severity.

**Conclusion:** Established facial and vocal measurements, collected remotely in schizophrenia patients via smartphones in response to automated task prompts, demonstrated accuracy as markers of schizophrenia severity. Clinical implications are discussed.

## 1 Introduction

There is rapidly increasing utilization of remote digital measurements in clinical research and practice. The development and validation of digital measurement tools in psychiatry come with both significant opportunities and risks. Significant opportunity arises as psychiatry is already undergoing a paradigm shift towards the utilization of objective markers to assess illness and disease progression (Insel, 2018) and towards the widespread use of telehealth platforms for psychiatric care. This is particularly important when face-to-face medical care is not possible, such as during the COVID-19 pandemic (Figueroa & Aguilera, 2020).

Many behavioral and physiological markers are now accessible through digital technology such as wearables, mobile/web apps, and application programming interfaces (APIs; Insel, 2017). Such advances hold promise in allowing new innovations in neuropsychiatry to truly scale in a manner where they can be utilized to develop and implement treatment for patients suffering from significant psychiatric impairment (Insel, 2019).

Schizophrenia represents a poignant example of both the benefits and challenges of remote digital measurement. Clinical trials for schizophrenia drug development are often site-centric, requiring patients to appear physically at the site for measurement of disease severity. The need to travel to sites can restrict study populations to those that live in geographical proximity to the site, restricting access to participation and limiting patient diversity (Lecomte et al., 2020). Current approaches for measurement of disease rely on clinician administered measures that are costly and time-consuming to administer, leading to infrequent assessment and low adherence to treatment regimens. The instruments themselves are not well-aligned with current neurobiological definitions of illness (Torous et al., 2018). For example, cognitive and motor dysfunction, which are symptoms of the negative subtype of schizophrenia, are not accurately assessed through existing scales.

Remote digital measurements are not prone to the same level of subjectivity as human raters, and can provide both scale and ease of use while better aligning clinical measurement with behavioral and physiological measurements of underlying neurobiological treatment targets (Marsch, 2018; Torous & Keshavan, 2018). Indeed, a large number of physiological and behavioral measures have demonstrated validity as markers of schizophrenia severity or caseness. Such markers, often examined in controlled laboratory settings, hold promise as accessible remote proxies to track clinical functioning in patients with schizophrenia. However, there is a need to determine their reliability when captured in real world settings, where differentiating between significant variability and noise can pose a challenge.

A number of behavioral characteristics of schizophrenia, such as alogia (poverty of speech) and affective flattening (diminished emotional expression/emotional withdrawal; (Tandon et al., 2013) can be quantified directly using standardized tasks and coding schemes (Alberto et al., 2019; de Boer et al., 2020; Kohler, Martin, Milonova, et al., 2008; Mandal et al., 1998; Mattes et al., 1995), which can be automated through use of computer vision (Baltrusaitis et al., 2016) and vocal acoustic (Jadoul et al., 2018) machine learning models. In addition to digital measures that are directly analogous to core schizophrenia symptomatology, there are a number of other acoustic measures including vocal loudness, pitch variability, fundamental frequency, and jitter that have demonstrated validity as markers of schizophrenia (Alberto et al., 2019; Covington et al., 2012; Martínez-Sánchez et al., 2015; Saxman & Burk, 1968). These markers have demonstrated specificity as measures of the negative symptom cluster in particular (Covington et al., 2012).

In the current investigation, we examine the ability to measure schizophrenia severity through facial and vocal analysis using videos recorded during a remote smartphone-based assessment composed of both evoked and spontaneous prompts. We compare these measures against standard clinical assessments of overall schizophrenia severity (i.e. Positive and Negative Syndrome Scale (PANSS) Total) as well as specific domains of positive (P Total), negative (N Total), and general (G Total) symptoms, measured during clinic visits. We further examine the relationship between digital measures and individual symptoms of schizophrenia.

## 2 Methods

### 2.1 Participants

Individuals who passed a phone-screen for a DSM-5 diagnosis of schizophrenia and were on a stable treatment regimen for atypical antipsychotic therapy for two months or more with no intent to change medication during the two-week study were recruited as study participants. A total of 20 individuals with schizophrenia were enrolled (8 males, 12 females) with an age range of 29 to 61 (*µ* = 45, σ = 11). To be included in the study, participants needed to have the ability to be able to speak, read, hear, and understand the language of the clinical staff and the Informed Consent Form, respond verbally to questions, follow instructions, and be willing and be able to participate in all study activities, including the use of smartphones for data collection as described in Section 2.2. The study was conducted at the Mount Sinai Health System Outpatient Psychiatry Clinics and the protocol was approved by the Biomedical Research Alliance of New York (BRANY).

### 2.2 Data collection

All study participants were assessed for severity of schizophrenia symptomatology using both in-person clinical assessments and remote smartphone-based assessments over the course of the 14-day observational period.

### 2.2.1 In-person clinical assessments

The Positive and Negative Syndrome Scale (PANSS) was administered in person for all participants by clinic staff on the first (day 1) and last (day 14) of the study. For all subsequent analyses, the PANSS scores for each study participant were averaged for the two time points. Given the study participants were clinically stable, averaging the two PANSS scores allowed for reduction in any noise in the measurement. In addition to the PANSS, all participants were assessed for negative symptomatology, with a binary yes/no determination of whether or not they were demonstrating negative symptoms of schizophrenia.

### 2.2.2 Remote smartphone-based assessments

On the first day of the study, all study participants were trained by clinic staff on how to use the smartphone application (www.aicure.com) for remote data collection. The smartphone application was used to participate in remote assessments that would capture video and audio of participant behavior using the smartphone front-facing camera as they responded to on-screen prompts (Figure 1). Participants were allowed to use their own smartphones or use smartphones provisioned to them by the clinic staff for the duration of the study. The assessments were taken at scheduled time points over the course of the 14 days and were designed to capture two main kinds of behaviors as described below.

**Figure 1:**
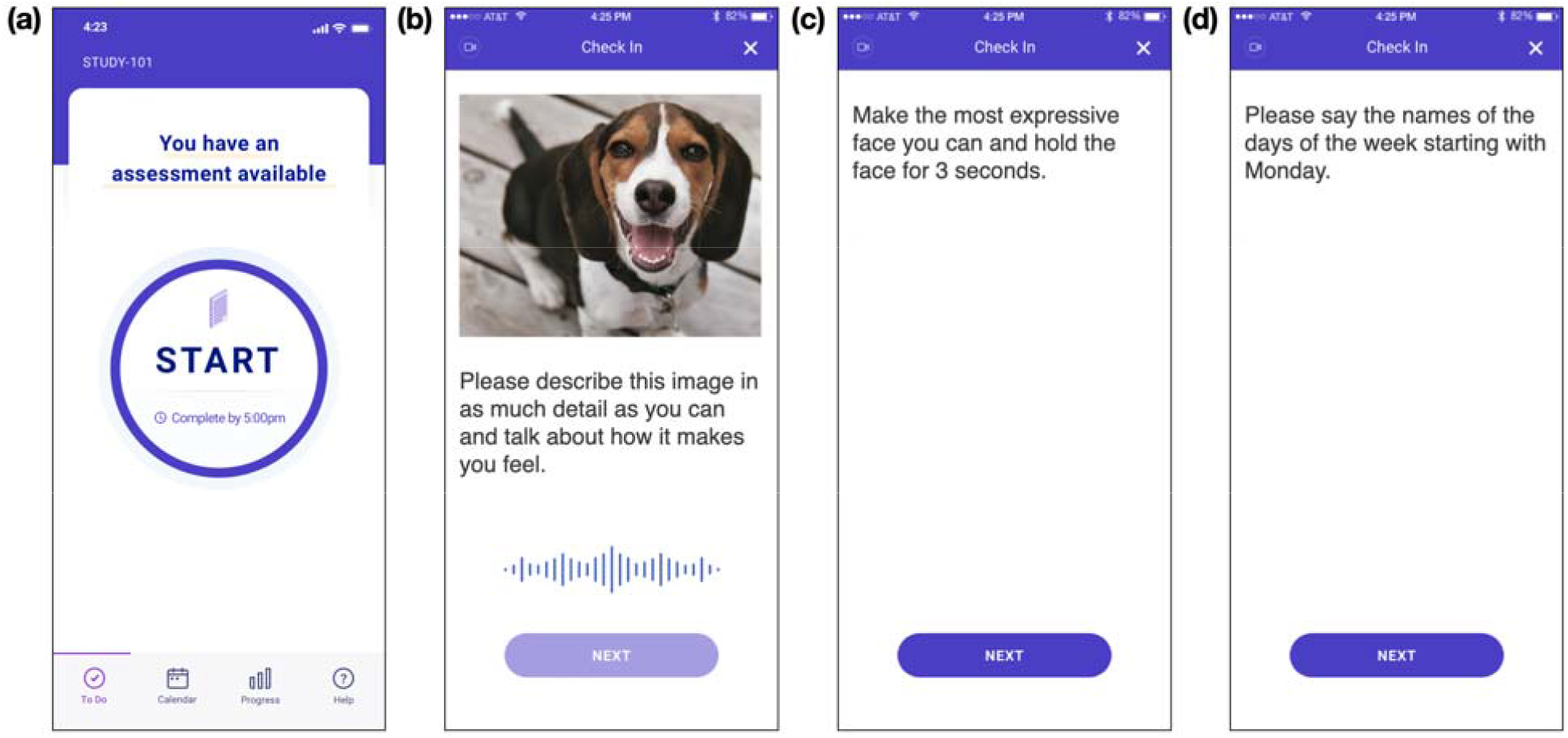
Example screenshots from the smartphone assessment all study participants took for remote and automated collection of video and audio data. During each of the prompts, the app speaks the text displayed on the screen and awaits a verbal and visual response from the participant, all while recording video and audio from the front-facing camera and microphone. **(a)** Screen displayed before the participant begins the assessment. **(b)** Prompt for collection of free behavior in response to images, showing one example image. **(c)** Prompt for collection of evoked facial expression behavior. **(d)** Prompt for collection of evoked vocal expression behavior.

#### Free speech and spontaneous expressivity

Participants were shown images from the Open Affective Standard Image Set (Kurdi et al., 2017) and asked to describe the images and talk about how they made them feel (Figure 1b). The participants’ speech and facial expressivity in response to the prompts were captured (Alberto et al., 2019; Cohen et al., 2016; Kohler, Martin, Milonova, et al., 2008; Kohler, Martin, Stolar, et al., 2008; Mandal et al., 1998; Mattes et al., 1995; Schwartz et al., 2006). This assessment was conducted on days 2, 7, and 14 of the study. Measurements acquired from each timepoint of the assessments were averaged for reduction of noise before comparison with PANSS.

#### Evoked facial and vocal expressions

Participants were asked separately to make the most expressive face they could and hold it for 3 seconds (Figure 1c) and then say the names of the days of the week out loud (Figure 1d). These prompts were selected based on prior experimental tasks used to examine emotional activity and speech in schizophrenia (Alpert et al., 1997; Kohler, Martin, Stolar, et al., 2008). The captured video and audio was used to measure facial expressivity and acoustic characteristics of voice during the evoked expressions. These assessments were scheduled on days 1, 7, and 14 of the study. Measurements across the time points were averaged for reduction of noise before comparison with PANSS.

### 2.3 Measurement of digital markers

Video and audio of participant behavior collected during the remote smartphone assessments was securely uploaded for processing. A combination of computer vision and digital signal processing tools were used for quantification of facial and vocal behavior and subsequent derivation of visual and auditory markers of schizophrenia as described below. The code with which these measures were acquired have been packaged as an open source software library and made available for use to all researchers on Github: https://github.com/aicure/open_dbm.

#### 2.3.1 Measurement of facial expressivity

The software library OpenFace (https://github.com/TadasBaltrusaitis/OpenFace) was used to measure framewise facial expressivity through quantification of action units (AUs; Supplementary Table 1) using a computer vision-based implementation of the Facial Action Coding System (FACS). All framewise AU measurements were normalized through division by a timepoint-specific baseline value acquired at the beginning of each assessment when the participant is not presented with any stimulus. The normalization allows for correction of any inter- and intra-individual variability; this methodology has previously been demonstrated to be necessary for measurement of facial behavior using computer vision tools and for subsequent analyses of facial expressivity (Alvino et al., 2007; Wang et al., 2007; Wang et al., 2008). *Facial expressivity* was calculated by taking the mean framewise intensity of all AUs over the course of the video. The method for quantifying *facial expressivity* was the same for both spontaneous and evoked expressivity. For each frame of video, OpenFace provides a confidence score denoting the likelihood that it is accurately detecting a face; only frames with a confidence score of 80% or higher were used for all downstream analysis. While OpenFace provides large amounts of information on specific AUs and emotions, in the current investigation, we focused only on *facial expressivity* because of significant evidence that patients with schizophrenia display less overall affect (e.g. blunted affect; (Berenbaum & Oltmanns, 1992; Henry et al., 2007).

#### 2.3.2 Measurement of vocal acoustics

The software library Parselmouth (https://github.com/YannickJadoul/Parselmouth), which is a python implementation of the Praat software library (www.praat.org), was used for measurement of all vocal acoustic characteristics. All audio analyzed was first passed through the LogMMSE noise-reduction algorithm for speech enhancement (Cannizzaro et al., 2005; Jadoul et al., 2018). Measurements of the verbal acoustic features listed in Table 1 were extracted from the audio collected. Each of the features were calculated separately during free speech and evoked vocal expression.

**Table 1:** List of vocal acoustic variables extracted from audio files collected during participation in remote smartphone assessments.

| Variable | Description |
| --- | --- |
| *Vocal intensity* | Volume of participant's speech, measured in dB |
| *Fundamental frequency mean* | Average fundamental frequency of participant speech in Hz |
| *Fundamental frequency stdev* | Standard deviation in fundamental frequency in Hz |
| *Vocal jitter* | Degree of irregularity in the frequency of the participant's speech, measured in Hz |
| *Speech prevalence* | Percentage of the audio file where participant speech was detected as opposed to silence |
| *Harmonics to noise ratio* | Quantification of additive noise in the participant's speech |

**Table 2:** All variables described in Section 2.3 were calculated separately for distinct behaviors captured during the remote smartphone assessments. Each of the behaviors that were elicited and captured during the smartphone assessment and the digital markers calculated from those behaviors are listed here.

| Behavior | On-screen prompt | Digital markers measured |
| --- | --- | --- |
| Free behavior | *Please describe what you see in this image and talk about how it makes you feel* (Figure 1b) | *Facial expressivity*<br>*Fundamental frequency mean*<br>*Fundamental frequency stdev*<br>*Vocal Jitter*<br>*Harmonics-to-noise ratio*<br>*Speech prevalence* |
| Evoked facial expression | *Please make the most expressive face you can and hold it for 3 seconds* (Figure 1c) | *Facial expressivity* |
| Evoked vocal expression | Please say the names of the days of the week starting with Monday (Figure 1d) | Fundamental frequency mean<br>Fundamental frequency stdev<br>Vocal Jitter<br>Harmonics-to-noise ratio<br>Speech prevalence |

Despite the exploratory nature of this study and given the small data sample, we attempted to be parsimonious in selection of markers to reduce the likelihood of false discovery. Analysis of vocal markers included those that have previously demonstrated effects in studies of individuals with schizophrenia (Alberto et al., 2019; Martínez-Sánchez et al., 2015). These properties, recorded both during free speech and evoked vocal expressions, include loudness of the individual’s voice in Decibels (*vocal intensity*), average fundamental frequency in Hertz (*fundamental frequency mean*), standard deviation of fundamental frequency (*fundamental frequency stdev*), jitter (*vocal jitter*), harmonics-to-noise ratio (*harmonics to noise ratio*) and the percentage of time with detected speech in an audio file (*speech prevalence)* (Cannizzaro et al., 2005; Covington et al., 2012; Kliper et al., 2019; Sarioglu Kayi et al., 2017; Saxman & Burk, 1968).

### 2.4 Data analysis

Both facial expressivity and vocal characteristics were assessed during free behavior following spontaneous prompts. Facial expressivity was also assessed during evoked facial expressions and vocal characteristics were assessed during evoked vocal expression following evoked prompts. Evaluation of vocal characteristics during the evoked expression task allowed for measurement of specific characteristics that have been previously shown to be effective measures of schizophrenia during phonation (e.g. *fundamental frequency mean and stdev, jitter, harmonics-to-noise ratio*) while also measuring speech characteristics such as amount of time spoken (i.e. *speech prevalence*) (Cannizzaro et al., 2005; Covington et al., 2012; Kliper et al., 2019; Sarioglu Kayi et al., 2017; Saxman & Burk, 1968). A large number of variables can be calculated from video and audio data sources; however, the analyses presented herein were limited to features that have evidence and a theoretical basis for relationship to schizophrenia severity and symptoms in the scientific literature.

#### 2.4.1 Comparison to PANSS subscale scores

Digital measures were compared to schizophrenia severity overall using the PANSS total severity score (*PANSS Total*) along with the three subscales reflecting negative symptom severity (*N Total*), positive symptom severity (*P Total*), and general severity (*G Tota*l) using Pearson’s correlation. When comparing negative symptoms, we utilized the PANSS Marder Symptom Factor which includes two symptoms that are traditionally included in the general severity score: *Motor Retardation* and *Social Avoidance and Isolation*.

#### 2.4.2 Comparison to individual PANSS items

Digital measurements that demonstrated significance in relation to specific subscales were then further explored in relation to the specific symptoms that derive those subscales, correcting for multiple comparisons using a Benjamini-Hochberg adjusted p-value (Li & Barber, 2019). This was an exploratory analysis conducted to further disaggregate the heterogeneity within the symptom scales to understand more specifically which clinical features were reflected in the digital measurement.

## 3 Results

### 3.1 Comparison to PANSS scores

#### 3.1.1 Vocal markers during evoked vocal expression

Results demonstrate that multiple digital measures are significantly correlated with overall negative symptom severity (N Total) after correcting for multiple comparisons. This includes *fundamental frequency mean* (*r* = -0.64; *adjusted p* = .02), *vocal jitter* (*r* = 0.56; *adjusted p* = .02), and *harmonics to noise ratio* (*r* = -0.61; *adjusted p* = .02). Two other features demonstrated marginal significance after correction for false discovery, including *speech prevalence* (*r* = -0.47; *adjusted p* = .06) and *fundamental frequency stdev* (*r* = -0.44; *adjusted p* = .07; see Table 3 for full results). Importantly, the directionality of results was consistent with prior research. For example, increased negative symptom severity was reflected in decreased speech prevalence, decreased tonal qualities of speech, and increased noise to speech sounds, consistent with the literature (Alberto et al., 2019; Covington et al., 2012; Martínez-Sánchez et al., 2015; Saxman & Burk, 1968).

**Table 3:** Correlation between vocal markers during evoked vocal expression and PANSS score showed a relationship between vocal characteristics and schizophrenia severity.

| Variable |  | N Total | P Total | G Total | Total | Vocal intensity | Fundamental frequency stdev | Fundamental frequency mean | Vocal jitter | Speech prevalence |
| --- | --- | --- | --- | --- | --- | --- | --- | --- | --- | --- |
| 1. N Total | Pearson's r<br>p-value | —<br>— |  |  |  |  |  |  |  |  |
| 2. P Total | Pearson's r<br>p-value | 0.452*<br>0.045 | —<br>— |  |  |  |  |  |  |  |
| 3. G Total | Pearson's r<br>p-value | 0.572**<br>0.008 | 0.806***<br>< .001 | —<br>— |  |  |  |  |  |  |
| 4. Total | Pearson's r<br>p-value | 0.757***<br>< .001 | 0.870***<br>< .001 | 0.947***<br>< .001 | —<br>— |  |  |  |  |  |
| 5. Vocal intensity | Pearson's r<br>p-value | -0.091<br>0.710 | -0.250<br>0.903 | -0.088<br>0.720 | -0.152<br>0.642 | —<br>— |  |  |  |  |
| 6. Fundamental frequency stdev | Pearson's r<br>p-value | -0.436<br>0.074 | -0.068<br>0.782 | 0.098<br>0.827 | -0.090<br>0.714 | -0.081<br>0.743 | —<br>— |  |  |  |
| 7. Fundamental frequency mean | Pearson's r<br>p-value | -0.644*<br>0.018 | -0.253<br>0.296 | -0.218<br>0.371 | -0.373<br>0.696 | 0.475<br>0.1 | 0.577*<br>0.020 | —<br>— |  |  |
| 8. Vocal jitter | Pearson's r<br>p-value | 0.563*<br>0.024 | 0.229<br>0.519 | 0.122<br>0.928 | 0.293<br>0.336 | -0.176<br>0.786 | -0.695***<br>< .001 | -0.823***<br>< .001 | —<br>— |  |
| 9. Speech prevalence | Pearson's r<br>p-value | -0.470<br>0.063 | -0.247<br>0.614 | -0.292<br>0.225 | -0.362<br>0.381 | 0.611*<br>0.025 | 0.043<br>0.863 | 0.781***<br>< .001 | -0.373<br>0.116 | —<br>— |
| 10. Harmonics to noise ratio | Pearson's r<br>p-value | -0.610*<br>0.018 | -0.195<br>0.507 | -0.126<br>0.606 | -0.297<br>0.434 | 0.154<br>0.66 | 0.773***<br>< .001 | 0.868***<br>< .001 | -0.965***<br>< .001 | 0.422<br>0.072 |

\* $p < 0.05$ ; \*\* $p < 0.01$ ; \*\*\* $p < 0.001$

Upon examination of the relationship between vocal measures and individual negative symptoms, results demonstrated that specific symptoms including *Emotional Withdrawal (r = - 0*.*55* ; *p =* .*015), Poor Rapport (r = -0*.*59* ; *p =* .*008), Lack of Spontaneity (r = -0*.*59; p =* .*008*, and *Motor Retardation (r = -0*.*53; p =* .*02)* demonstrated significant correlations with multiple auditory measures in a direction consistent with their relationship to overall negative symptom severity (Supplementary Table 2).

#### 3.1.2 Evoked facial expression

Results demonstrated that *facial expressivity* demonstrated significant relationships with overall schizophrenia severity PANSS Total (*r* = -0.71; *adjusted p* = .002) and severity on all PANSS subscales (N Total, *r* = -0.50; *adjusted p* = .035; P Total, *r* = -0.63; *adjusted p* = .006; G Total, *r* = -0.70; *adjusted p* = .009). See Table 4.

**Table 4:**
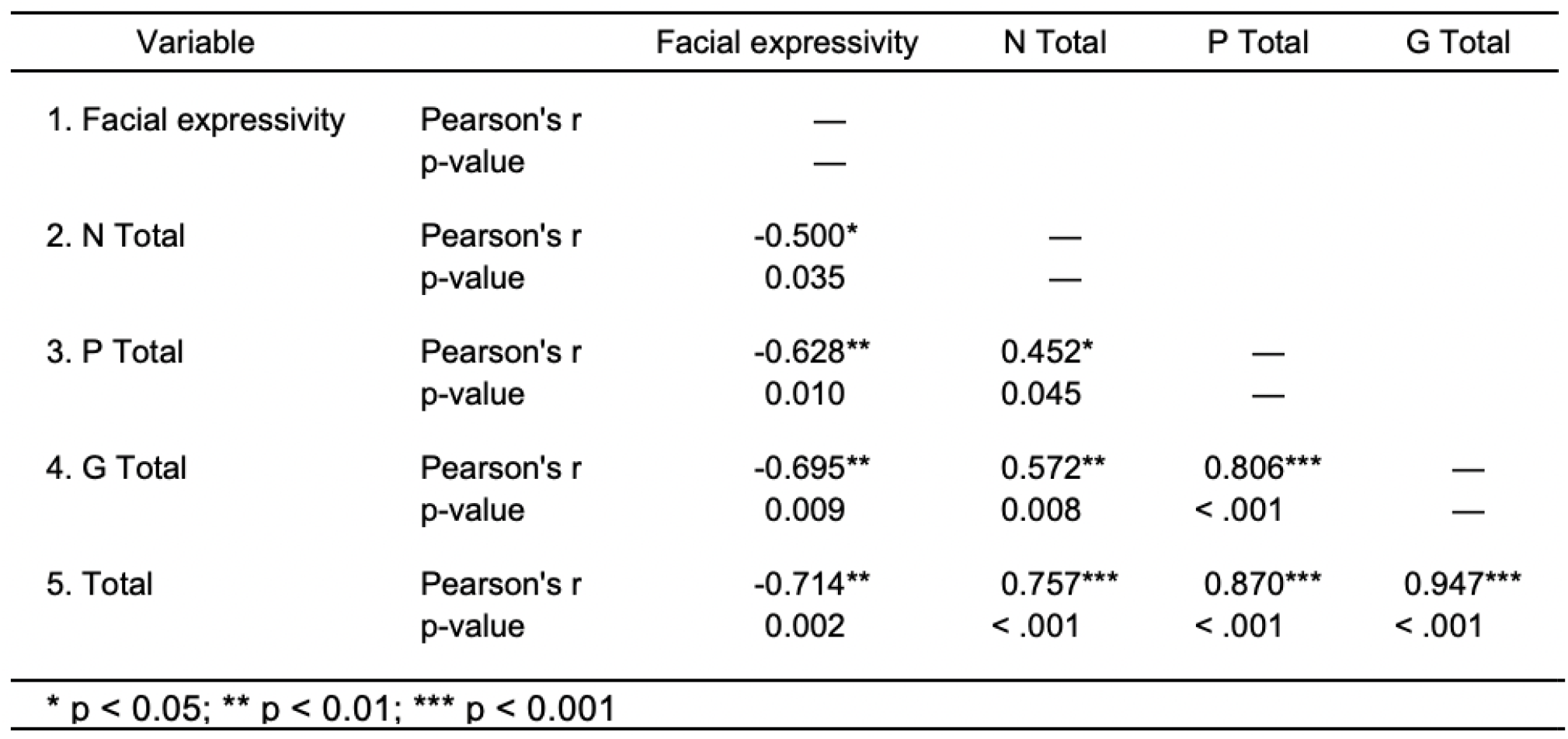
Correlation between *facial expressivity* during evoked facial expression and PANSS score showed a relationship between facial affect and schizophrenia severity.

Pearson’s correlations with individual symptoms demonstrates significant or marginally significant relationships with multiple individual negative symptoms including *Blunted Affect* (*r* = -0.49; *p* = .04), *Social Withdrawal* (*r* = -0.63; *p* = .005), *Stereotyped Thinking* (*r* = -0.41; *p* = .09), and *Motor Retardation* (*r* = -0.45; *p* = .06). Further, facial expressivity was significantly negatively correlated with positive symptoms including *Delusions* (*r* = -0.81; *p* < .001), *Hallucinations* (*r* = -0.50; *p* = .04), and *Paranoid Ideation* (*r* = -0.64; *p* = .005). Finally, *facial expressivity* demonstrated significant negative correlations with symptoms in the general symptom factor including *Guilt* (*r* = -0.50; *p* = .035), *Tension* (*r* = -0.40; *p* = .10), *Unusual Thought Content* (*r* = -0.65; *p* = .02), *Disturbances in Volition* (*r* = -0.78; *p* < .001), and *Social Avoidance* (*r* = -0.79; *p* < .001). See Supplementary Table 3 for complete results.

#### 3.1.3 Free behavior in response to images

Spontaneous measurement of voice and facial expressions, as elicited by emotionally valenced images, demonstrated relationships between multiple vocal markers and the negative symptom cluster. Highly consistent with results of vocal measurements in response to evoked prompts, the following measures demonstrated significance in relation to negative symptom severity (N Tota)l: *fundamental frequency mean* (*r* = -0.61; *adjusted p* = .04), *harmonics to noise ratio* (*r* = - 0.58; *adjusted p* = .03), *speech prevalence* (*r* = -0.57; *adjusted p* = .025). *Vocal jitter* demonstrated a marginally significant adjusted p-value (*r* = 0.43; *adjusted p* = .09), and *fundamental frequency stdev* did not approach significance (See Table 5). In contrast to measurement after the evoked task, *vocal intensity* measured during free behavior did demonstrate significance (*r* = 0.50; *adjusted p* = .05).

**Table 5:** Correlation between facial and vocal markers during free behavior and PANSS score showed a relationship between facial affect and vocal characteristics with schizophrenia severity.

| Variable |  | N Total | P Total | G Total | Total | Facial expressivity | Vocal intensity | Fundamental frequency mean | Fundamental frequency Stdev | Harmonics to noise ratio | Vocal jitter |
| --- | --- | --- | --- | --- | --- | --- | --- | --- | --- | --- | --- |
| 1. N Total | Pearson's r | — |  |  |  |  |  |  |  |  |  |
|  | p-value | — |  |  |  |  |  |  |  |  |  |
| 2. P Total | Pearson's r | 0.452* | — |  |  |  |  |  |  |  |  |
|  | p-value | 0.045 | — |  |  |  |  |  |  |  |  |
| 3. G Total | Pearson's r | 0.572** | 0.806*** | — |  |  |  |  |  |  |  |
|  | p-value | 0.008 | < .001 | — |  |  |  |  |  |  |  |
| 4. Total | Pearson's r | 0.757*** | 0.870*** | 0.947*** | — |  |  |  |  |  |  |
|  | p-value | < .001 | < .001 | < .001 | — |  |  |  |  |  |  |
| 5. Facial expressivity | Pearson's r | 0.142 | -0.113 | 0.090 | 0.056 | — |  |  |  |  |  |
|  | p-value | 0.583 | 0.644 | 0.834 | 0.819 | — |  |  |  |  |  |
| 6. Vocal intensity | Pearson's r | -0.502* | -0.332 | -0.225 | -0.386 | 0.364 | — |  |  |  |  |
|  | p-value | 0.050 | 0.165 | 0.826 | 0.240 | 0.126 | — |  |  |  |  |
| 7. Fundamental frequency mean | Pearson's r | -0.606* | -0.288 | -0.268 | -0.428 | 0.184 | 0.935*** | — |  |  |  |
|  | p-value | 0.042 | 0.812 | 0.268 | 0.476 | 0.451 | < .001 | — |  |  |  |
| 8. Fundamental frequency stdev | Pearson's r | -0.304 | -0.189 | -0.127 | -0.225 | 0.179 | 0.581** | 0.529* | — |  |  |
|  | p-value | 0.240 | 0.613 | 0.605 | 0.495 | 0.464 | 0.009 | 0.020 | — |  |  |
| 9. Harmonics to noise ratio | Pearson's r | -0.584* | -0.224 | -0.097 | -0.312 | 0.174 | 0.654** | 0.774*** | 0.476* | — |  |
|  | p-value | 0.031 | 0.624 | 0.970 | 0.339 | 0.475 | 0.002 | < .001 | 0.039 | — |  |
| 10. Vocal jitter | Pearson's r | 0.426 | 0.147 | 0.015 | 0.194 | -0.097 | -0.541* | -0.691** | -0.278 | -0.937*** | — |
|  | p-value | 0.096 | 0.640 | 0.951 | 0.498 | 0.692 | 0.017 | 0.001 | 0.249 | < .001 | — |
| 11. Speech prevalence | Pearson's r | -0.567* | -0.260 | -0.261 | -0.403 | 0.161 | 0.869*** | 0.923*** | 0.260 | 0.575** | -0.510* |
|  | p-value | 0.025 | 0.660 | 0.980 | 0.304 | 0.510 | < .001 | < .001 | 0.283 | 0.010 | 0.026 |

\* $p < 0.05$ ; \*\* $p < 0.01$ ; \*\*\* $p < 0.001$

Individual negative symptoms broadly demonstrated relationships with multiple vocal markers (See Supplementary Table 4 for complete results). *Blunted affect* only demonstrated a marginally significant relationship with *speech prevalence* (*r* = 0.40; *p* = .09) while *vocal intensity* only demonstrated a significant relationship with the symptom of *Emotional Withdrawal* (*r* = - 0.53; *p* = .02).

## 4 Discussion

In the current investigation, we sought to test the hypothesis that facial and vocal markers of schizophrenia can be captured remotely in patients using brief automated smartphone-based assessments and that such measures would be well-correlated to standard clinical measures of schizophrenia symptom severity. Such measures show promise of objective and automated methods of assessing illness severity in the context of treatment development and decision making. Prompts and vocal/facial measures that have previously demonstrated accuracy in controlled research settings were simplified and deployed as a brief assessment via a smartphone application in an observational study with schizophrenia patients. Results support the ability to measure meaningful clinical markers of schizophrenia severity via a brief smartphone based assessment that captures data remotely and processes it through back-end deep machine learning algorithms to create vocal and facial markers.

Results demonstrate that vocal characteristics such as fundamental frequency, loudness, non-verbal vocal tones and prevalence of speech serve as specific markers of negative symptom severity. The majority of these markers demonstrate a robust signal of negative symptom severity regardless of whether prompts were evoked or spontaneous.

The observation that vocal markers provide specificity as a metric of negative symptom severity has significant practical implications for clinical research and decision making. Recent advances in the mechanistic understanding of negative symptomatology have led to a number of promising pharmacological and cognitive treatments for negative symptoms of schizophrenia (Erhart et al., 2006; Fusar-Poli et al., 2015; Millan et al., 2014; Singh et al., 2010). Such initiatives are important given the lack of FDA-approved treatments for negative symptoms (Kirkpatrick et al., 2006). However, reliable and change-sensitive measures of negative symptomatology to assess the efficacy of these treatments are sparse (King, 1998; Möller, 2007; Prikryl et al., 2007; Walther et al., 2009).

Facial expressivity only demonstrated a relationship with schizophrenia severity when captured using evoked prompts. This may indicate that either greater structure is needed to assess this marker remotely or that the prompts that were utilized were not a strong enough elicitation. Indeed, prior work has demonstrated that video rather than still images are stronger evocations to assess emotional variability in schizophrenia (Bersani et al., 2013). Despite this, we do observe that facial expressivity in response to evoked prompts provides a robust signal for overall symptom severity. Analysis of single symptoms demonstrates face validity for this marker as it reduced facial expressivity relates to greater blunted affect, social withdrawal, social avoidance, and motor retardation. We also observe that facial expressivity was decreased as subjects endorsed greater positive symptoms of hallucinations, delusions, and paranoid ideation. This finding is consistent with evidence that social and cognitive impairment, which falls into the negative symptom cluster, mechanistically relates to neurodegeneration that also impacts visual and auditory hallucinations (Bersani et al., 2013; Jenkins et al., 2018; Zhuo et al., 2020).

The current study presents with a number of important limitations. While the primary hypotheses were supported, not all effects were consistent across prompts. Given the small sample size, it is impossible to conclude definitively which markers can be utilized to robustly assess schizophrenia symptom severity or impairment. Indeed, a number of relatively large correlation coefficients demonstrated only marginal significance, likely due to sample size constraints. Further, despite the markers being hypothesized a priori, the current work is exploratory in nature given the small sample size, limited number of assessments, and the short duration of the study. A larger assessment will be needed to replicate the current findings. Despite the above limitations, the current work provides evidence that facial and vocal digital measures can be remotely captured in schizophrenia patients and that such measures demonstrate statistically significant relationships with established measures of schizophrenia symptom severity, demonstrating promise that these tools could be used to remotely measure and track schizophrenia symptoms and severity in an objective manner.

Finally, while the app-based video/audio capture utilizes a proprietary platform, this investigation utilized open-source Python-based software, available to all researchers (https://github.com/AiCure/open_dbm/). As an additional measure, the code that implemented the open-source software for this investigation and subsequent analyses of results have been provided by the authors in the Methods section. This allows for the expansion of the experiment to a wider patient population as mentioned above and the independent validation of the methods and their implementation in this investigation by other researchers in academic and clinical research, following an open-science framework for the development of digital tools for objective, accurate, and scalable measurement of disease symptomatology in both mental and physical health.

## 5 Conclusions

In this investigation, we demonstrate that facial and vocal markers, measured using computer vision and vocal analytics from video captured remotely via smartphones demonstrates validity as a marker of schizophrenia and is a promising metric for negative symptom severity. Use of such technology in clinical care and clinical research settings could allow for more frequent, remotely assessed, objective measurement of disease symptomatology and treatment response in a scalable and accessible manner, which can support development of novel treatments and risk assessment among individuals with schizophrenia.

## Data Availability

Due to privacy concerns, participants of this study did not agree for their data to be shared publicly.

## Acknowledgments

The authors appreciate the involvement of the clinical, research, and operations staff at both Mount Sinai and AiCure for the development, deployment, and implementation of the technology presented here and the participants who volunteered to be involved in the research.

## Supplementary Materials

**Supplementary Table 1:** List of facial action units (AUs) whose frame-wise intensity was quantified using computer vision; AU intensities were normalized and then combined to measure *facial expressivity*.

| Action Unit | Description |
| --- | --- |
| AU1 | Inner brow raiser |
| AU2 | Outer brow raiser |
| AU4 | Brow lowerer |
| AU5 | Upper lid raiser |
| AU6 | Cheek raiser |
| AU7 | Lid tightener |
| AU9 | Nose wrinkler |
| AU12 | Lip corner puller |
| AU15 | Lip corner depressor |
| AU16 | Lower lip depressor |
| AU20 | Lip stretcher |
| AU23 | Lip tightener |
| AU26 | Jaw drop |

**Supplementary Table 2:**
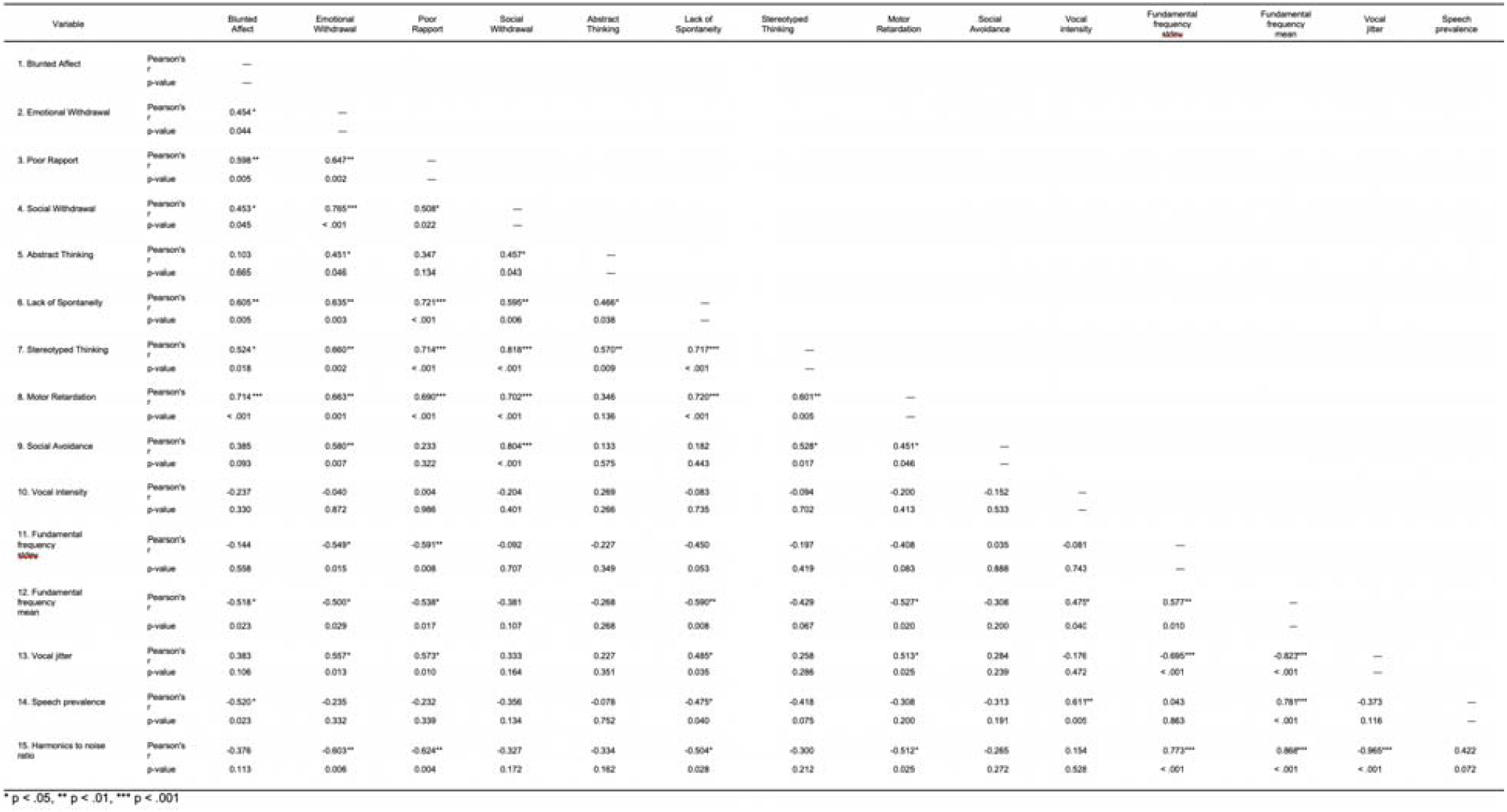
Correlation between vocal markers during evoked vocal expression and PANSS individual items.

**Supplementary Table 3:**
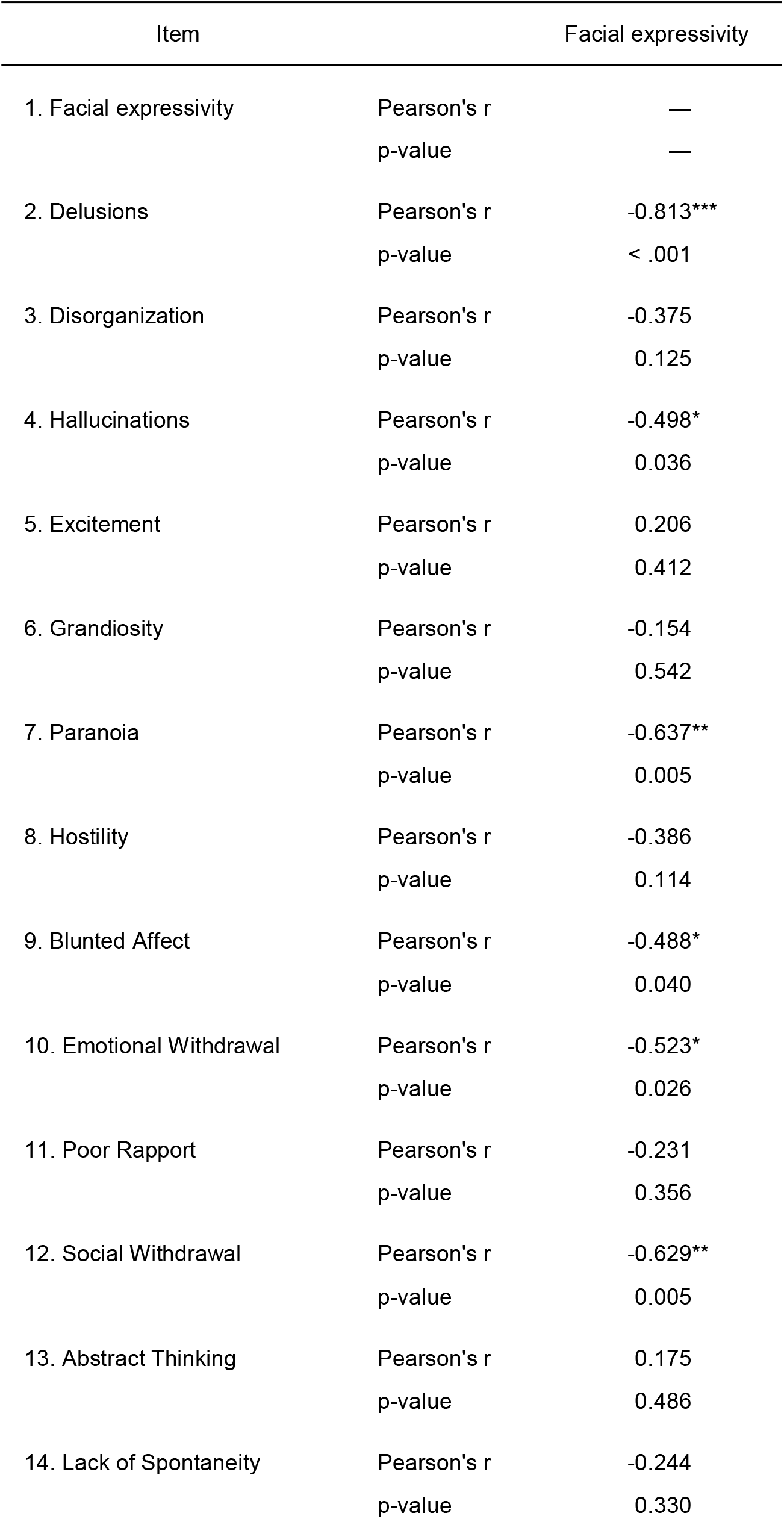
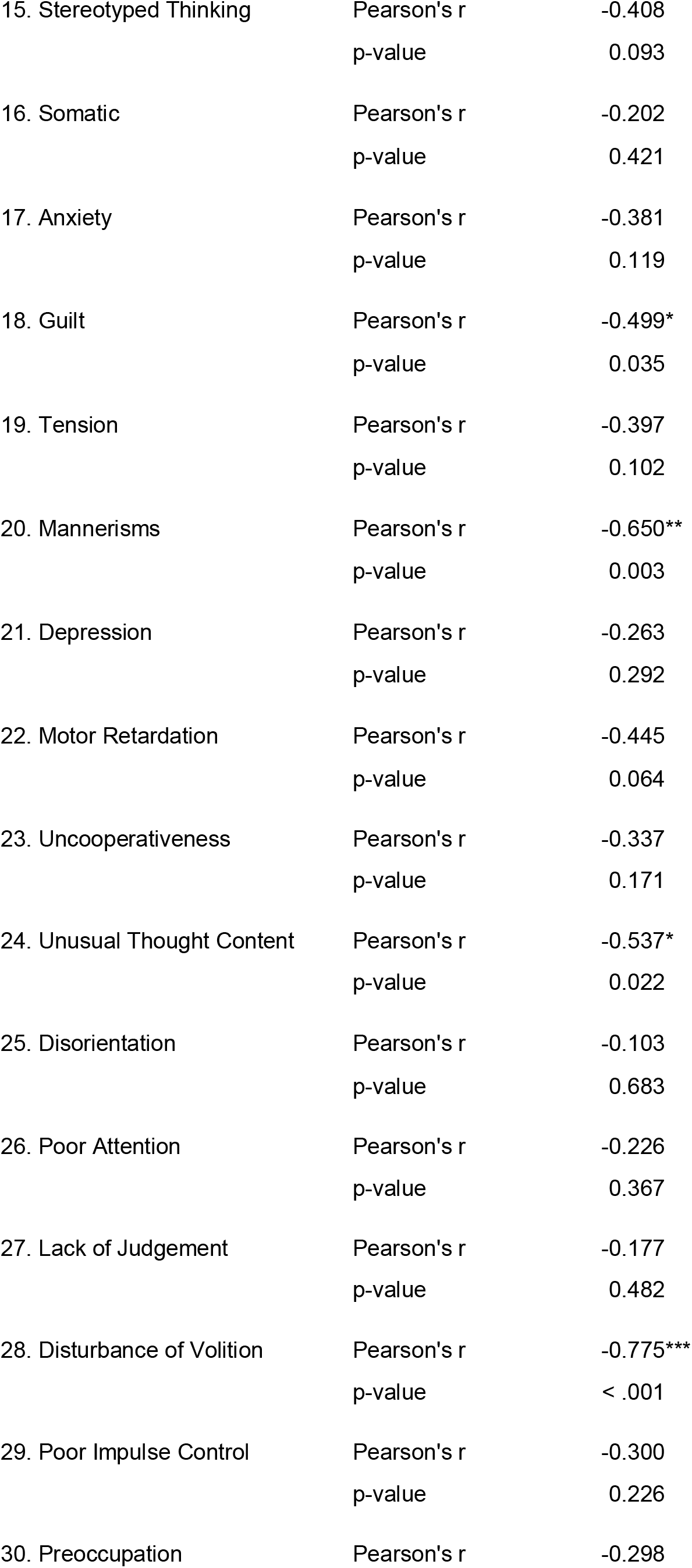
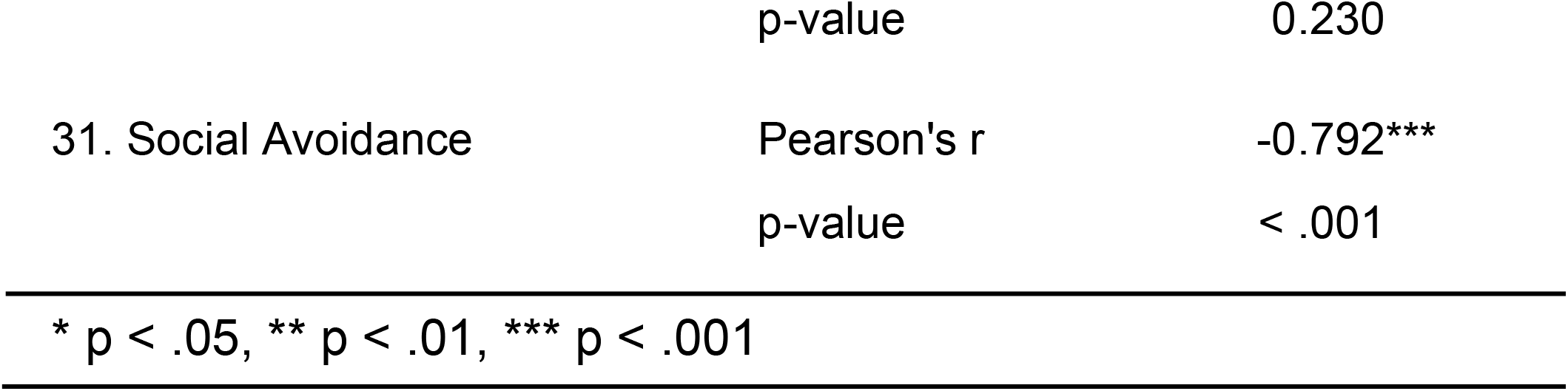
Correlation between facial markers during evoked facial expression and PANSS individual items.

**Supplementary Table 4:**
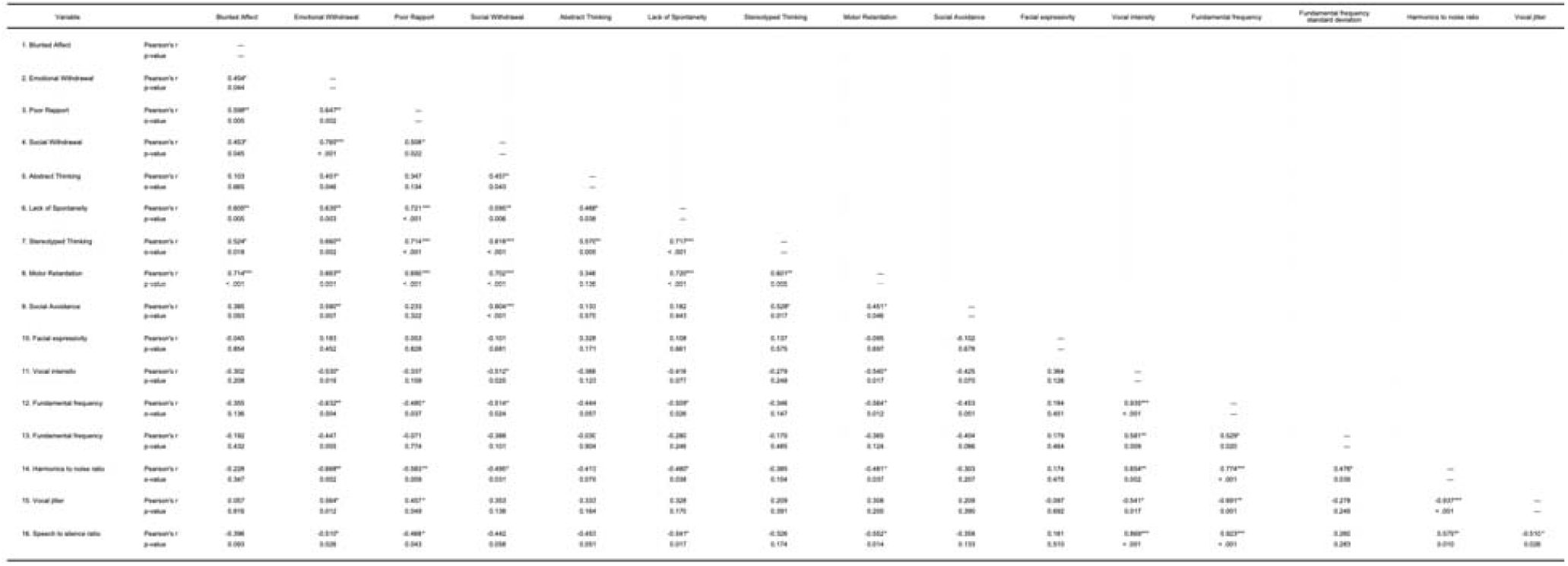
Correlation between facial and vocal markers during free behavior and PANSS individual items.

