## Supplemental Table 2 for "Facial and vocal markers of schizophrenia measured using remote smartphone assessments" (from another version of this preprint: version 1, 2020-12-04)

| Variable | |  | | Blunted Affect | | Emotional Withdrawal | | Poor Rapport | | Social Withdrawal | | Abstract Thinking | | Lack of Spontaneity | | Stereotyped Thinking | | Motor Retardation | | Social Avoidance | | Vocal intensity | | Fundamental frequency  stdev | | Fundamental frequency  mean | | Vocal jitter | | Speech prevalence |
| --- | --- | --- | --- | --- | --- | --- | --- | --- | --- | --- | --- | --- | --- | --- | --- | --- | --- | --- | --- | --- | --- | --- | --- | --- | --- | --- | --- | --- | --- | --- |
| 1. Blunted Affect |  | Pearson's r |  | — |  |  |  |  |  |  |  |  |  |  |  |  |  |  |  |  |  |  |  |  |  |  |  |  |  |  |
|  |  | p-value |  | — |  |  |  |  |  |  |  |  |  |  |  |  |  |  |  |  |  |  |  |  |  |  |  |  |  |  |
| 2. Emotional Withdrawal |  | Pearson's r |  | 0.454 | * | — |  |  |  |  |  |  |  |  |  |  |  |  |  |  |  |  |  |  |  |  |  |  |  |  |
|  |  | p-value |  | 0.044 |  | — |  |  |  |  |  |  |  |  |  |  |  |  |  |  |  |  |  |  |  |  |  |  |  |  |
| 3. Poor Rapport |  | Pearson's r |  | 0.598 | ** | 0.647 | ** | — |  |  |  |  |  |  |  |  |  |  |  |  |  |  |  |  |  |  |  |  |  |  |
|  |  | p-value |  | 0.005 |  | 0.002 |  | — |  |  |  |  |  |  |  |  |  |  |  |  |  |  |  |  |  |  |  |  |  |  |
| 4. Social Withdrawal |  | Pearson's r |  | 0.453 | * | 0.765 | *** | 0.508 | * | — |  |  |  |  |  |  |  |  |  |  |  |  |  |  |  |  |  |  |  |  |
|  |  | p-value |  | 0.045 |  | < .001 |  | 0.022 |  | — |  |  |  |  |  |  |  |  |  |  |  |  |  |  |  |  |  |  |  |  |
| 5. Abstract Thinking |  | Pearson's r |  | 0.103 |  | 0.451 | * | 0.347 |  | 0.457 | * | — |  |  |  |  |  |  |  |  |  |  |  |  |  |  |  |  |  |  |
|  |  | p-value |  | 0.665 |  | 0.046 |  | 0.134 |  | 0.043 |  | — |  |  |  |  |  |  |  |  |  |  |  |  |  |  |  |  |  |  |
| 6. Lack of Spontaneity |  | Pearson's r |  | 0.605 | ** | 0.635 | ** | 0.721 | *** | 0.595 | ** | 0.466 | * | — |  |  |  |  |  |  |  |  |  |  |  |  |  |  |  |  |
|  |  | p-value |  | 0.005 |  | 0.003 |  | < .001 |  | 0.006 |  | 0.038 |  | — |  |  |  |  |  |  |  |  |  |  |  |  |  |  |  |  |
| 7. Stereotyped Thinking |  | Pearson's r |  | 0.524 | * | 0.660 | ** | 0.714 | *** | 0.818 | *** | 0.570 | ** | 0.717 | *** | — |  |  |  |  |  |  |  |  |  |  |  |  |  |  |
|  |  | p-value |  | 0.018 |  | 0.002 |  | < .001 |  | < .001 |  | 0.009 |  | < .001 |  | — |  |  |  |  |  |  |  |  |  |  |  |  |  |  |
| 8. Motor Retardation |  | Pearson's r |  | 0.714 | *** | 0.663 | ** | 0.690 | *** | 0.702 | *** | 0.346 |  | 0.720 | *** | 0.601 | ** | — |  |  |  |  |  |  |  |  |  |  |  |  |
|  |  | p-value |  | < .001 |  | 0.001 |  | < .001 |  | < .001 |  | 0.136 |  | < .001 |  | 0.005 |  | — |  |  |  |  |  |  |  |  |  |  |  |  |
| 9. Social Avoidance |  | Pearson's r |  | 0.385 |  | 0.580 | ** | 0.233 |  | 0.804 | *** | 0.133 |  | 0.182 |  | 0.528 | * | 0.451 | * | — |  |  |  |  |  |  |  |  |  |  |
|  |  | p-value |  | 0.093 |  | 0.007 |  | 0.322 |  | < .001 |  | 0.575 |  | 0.443 |  | 0.017 |  | 0.046 |  | — |  |  |  |  |  |  |  |  |  |  |
| 10. Vocal intensity |  | Pearson's r |  | -0.237 |  | -0.040 |  | 0.004 |  | -0.204 |  | 0.269 |  | -0.083 |  | -0.094 |  | -0.200 |  | -0.152 |  | — |  |  |  |  |  |  |  |  |
|  |  | p-value |  | 0.330 |  | 0.872 |  | 0.986 |  | 0.401 |  | 0.266 |  | 0.735 |  | 0.702 |  | 0.413 |  | 0.533 |  | — |  |  |  |  |  |  |  |  |
| 11. Fundamental frequency  stdev |  | Pearson's r |  | -0.144 |  | -0.549 | * | -0.591 | ** | -0.092 |  | -0.227 |  | -0.450 |  | -0.197 |  | -0.408 |  | 0.035 |  | -0.081 |  | — |  |  |  |  |  |  |
|  |  | p-value |  | 0.558 |  | 0.015 |  | 0.008 |  | 0.707 |  | 0.349 |  | 0.053 |  | 0.419 |  | 0.083 |  | 0.888 |  | 0.743 |  | — |  |  |  |  |  |  |
| 12. Fundamental frequency  mean |  | Pearson's r |  | -0.518 | * | -0.500 | * | -0.538 | * | -0.381 |  | -0.268 |  | -0.590 | ** | -0.429 |  | -0.527 | * | -0.308 |  | 0.475 | * | 0.577 | ** | — |  |  |  |  |
|  |  | p-value |  | 0.023 |  | 0.029 |  | 0.017 |  | 0.107 |  | 0.268 |  | 0.008 |  | 0.067 |  | 0.020 |  | 0.200 |  | 0.040 |  | 0.010 |  | — |  |  |  |  |
| 13. Vocal jitter |  | Pearson's r |  | 0.383 |  | 0.557 | * | 0.573 | * | 0.333 |  | 0.227 |  | 0.485 | * | 0.258 |  | 0.513 | * | 0.284 |  | -0.176 |  | -0.695 | *** | -0.823 | *** | — |  |  |
|  |  | p-value |  | 0.106 |  | 0.013 |  | 0.010 |  | 0.164 |  | 0.351 |  | 0.035 |  | 0.286 |  | 0.025 |  | 0.239 |  | 0.472 |  | < .001 |  | < .001 |  | — |  |  |
| 14. Speech prevalence |  | Pearson's r |  | -0.520 | * | -0.235 |  | -0.232 |  | -0.356 |  | -0.078 |  | -0.475 | * | -0.418 |  | -0.308 |  | -0.313 |  | 0.611 | ** | 0.043 |  | 0.781 | *** | -0.373 |  | — |
|  |  | p-value |  | 0.023 |  | 0.332 |  | 0.339 |  | 0.134 |  | 0.752 |  | 0.040 |  | 0.075 |  | 0.200 |  | 0.191 |  | 0.005 |  | 0.863 |  | < .001 |  | 0.116 |  | — |
| 15. Harmonics to noise ratio |  | Pearson's r |  | -0.376 |  | -0.603 | ** | -0.624 | ** | -0.327 |  | -0.334 |  | -0.504 | * | -0.300 |  | -0.512 | * | -0.265 |  | 0.154 |  | 0.773 | *** | 0.868 | *** | -0.965 | *** | 0.422 |
|  |  | p-value |  | 0.113 |  | 0.006 |  | 0.004 |  | 0.172 |  | 0.162 |  | 0.028 |  | 0.212 |  | 0.025 |  | 0.272 |  | 0.528 |  | < .001 |  | < .001 |  | < .001 |  | 0.072 |

* p < .05, ** p < .01, *** p < .001
