## Supplemental Table 3 for "Facial and vocal markers of schizophrenia measured using remote smartphone assessments" (from another version of this preprint: version 1, 2020-12-04)

| Variable | |  | | Facial expressivity | |
| --- | --- | --- | --- | --- | --- |
| 1. Facial expressivity |  | Pearson's r |  | — |  |
|  |  | p-value |  | — |  |
| 2. Delusions |  | Pearson's r |  | -0.813 | *** |
|  |  | p-value |  | < .001 |  |
| 3. Disorganization |  | Pearson's r |  | -0.375 |  |
|  |  | p-value |  | 0.125 |  |
| 4. Hallucinations |  | Pearson's r |  | -0.498 | * |
|  |  | p-value |  | 0.036 |  |
| 5. Excitement |  | Pearson's r |  | 0.206 |  |
|  |  | p-value |  | 0.412 |  |
| 6. Grandiosity |  | Pearson's r |  | -0.154 |  |
|  |  | p-value |  | 0.542 |  |
| 7. Paranoia |  | Pearson's r |  | -0.637 | ** |
|  |  | p-value |  | 0.005 |  |
| 8. Hostility |  | Pearson's r |  | -0.386 |  |
|  |  | p-value |  | 0.114 |  |
| 9. Blunted Affect |  | Pearson's r |  | -0.488 | * |
|  |  | p-value |  | 0.040 |  |
| 10. Emotional Withdrawal |  | Pearson's r |  | -0.523 | * |
|  |  | p-value |  | 0.026 |  |
| 11. Poor Rapport |  | Pearson's r |  | -0.231 |  |
|  |  | p-value |  | 0.356 |  |
| 12. Social Withdrawal |  | Pearson's r |  | -0.629 | ** |
|  |  | p-value |  | 0.005 |  |
| 13. Abstract Thinking |  | Pearson's r |  | 0.175 |  |
|  |  | p-value |  | 0.486 |  |
| 14. Lack of Spontaneity |  | Pearson's r |  | -0.244 |  |
|  |  | p-value |  | 0.330 |  |
| 15. Stereotyped Thinking |  | Pearson's r |  | -0.408 |  |
|  |  | p-value |  | 0.093 |  |
| 16. Somatic |  | Pearson's r |  | -0.202 |  |
|  |  | p-value |  | 0.421 |  |
| 17. Anxiety |  | Pearson's r |  | -0.381 |  |
|  |  | p-value |  | 0.119 |  |
| 18. Guilt |  | Pearson's r |  | -0.499 | * |
|  |  | p-value |  | 0.035 |  |
| 19. Tension |  | Pearson's r |  | -0.397 |  |
|  |  | p-value |  | 0.102 |  |
| 20. Mannerisms |  | Pearson's r |  | -0.650 | ** |
|  |  | p-value |  | 0.003 |  |
| 21. Depression |  | Pearson's r |  | -0.263 |  |
|  |  | p-value |  | 0.292 |  |
| 22. Motor Retardation |  | Pearson's r |  | -0.445 |  |
|  |  | p-value |  | 0.064 |  |
| 23. Uncooperativeness |  | Pearson's r |  | -0.337 |  |
|  |  | p-value |  | 0.171 |  |
| 24. Unusual Thought Content |  | Pearson's r |  | -0.537 | * |
|  |  | p-value |  | 0.022 |  |
| 25. Disorientation |  | Pearson's r |  | -0.103 |  |
|  |  | p-value |  | 0.683 |  |
| 26. Poor Attention |  | Pearson's r |  | -0.226 |  |
|  |  | p-value |  | 0.367 |  |
| 27. Lack of Judgement |  | Pearson's r |  | -0.177 |  |
|  |  | p-value |  | 0.482 |  |
| 28. Disturbance of Volition |  | Pearson's r |  | -0.775 | *** |
|  |  | p-value |  | < .001 |  |
| 29. Poor Impulse Control |  | Pearson's r |  | -0.300 |  |
|  |  | p-value |  | 0.226 |  |
| 30. Preoccupation |  | Pearson's r |  | -0.298 |  |
|  |  | p-value |  | 0.230 |  |
| 31. Social Avoidance |  | Pearson's r |  | -0.792 | *** |
|  |  | p-value |  | < .001 |  |
