## Supplemental Table 4 for "Facial and vocal markers of schizophrenia measured using remote smartphone assessments" (from another version of this preprint: version 1, 2020-12-04)

| Variable | |  | | Blunted Affect | | Emotional Withdrawal | | Poor Rapport | | Social Withdrawal | | Abstract Thinking | | Lack of Spontaneity | | Stereotyped Thinking | | Motor Retardation | | Social Avoidance | | Facial expressivity | | Vocal intensity | | Fundamental frequency | | Fundamental frequency  standard deviation | | Harmonics to noise ratio | | Vocal jitter | |
| --- | --- | --- | --- | --- | --- | --- | --- | --- | --- | --- | --- | --- | --- | --- | --- | --- | --- | --- | --- | --- | --- | --- | --- | --- | --- | --- | --- | --- | --- | --- | --- | --- | --- |
| 1. Blunted Affect |  | Pearson's r |  | — |  |  |  |  |  |  |  |  |  |  |  |  |  |  |  |  |  |  |  |  |  |  |  |  |  |  |  |  |  |
|  |  | p-value |  | — |  |  |  |  |  |  |  |  |  |  |  |  |  |  |  |  |  |  |  |  |  |  |  |  |  |  |  |  |  |
| 2. Emotional Withdrawal |  | Pearson's r |  | 0.454 | * | — |  |  |  |  |  |  |  |  |  |  |  |  |  |  |  |  |  |  |  |  |  |  |  |  |  |  |  |
|  |  | p-value |  | 0.044 |  | — |  |  |  |  |  |  |  |  |  |  |  |  |  |  |  |  |  |  |  |  |  |  |  |  |  |  |  |
| 3. Poor Rapport |  | Pearson's r |  | 0.598 | ** | 0.647 | ** | — |  |  |  |  |  |  |  |  |  |  |  |  |  |  |  |  |  |  |  |  |  |  |  |  |  |
|  |  | p-value |  | 0.005 |  | 0.002 |  | — |  |  |  |  |  |  |  |  |  |  |  |  |  |  |  |  |  |  |  |  |  |  |  |  |  |
| 4. Social Withdrawal |  | Pearson's r |  | 0.453 | * | 0.765 | *** | 0.508 | * | — |  |  |  |  |  |  |  |  |  |  |  |  |  |  |  |  |  |  |  |  |  |  |  |
|  |  | p-value |  | 0.045 |  | < .001 |  | 0.022 |  | — |  |  |  |  |  |  |  |  |  |  |  |  |  |  |  |  |  |  |  |  |  |  |  |
| 5. Abstract Thinking |  | Pearson's r |  | 0.103 |  | 0.451 | * | 0.347 |  | 0.457 | * | — |  |  |  |  |  |  |  |  |  |  |  |  |  |  |  |  |  |  |  |  |  |
|  |  | p-value |  | 0.665 |  | 0.046 |  | 0.134 |  | 0.043 |  | — |  |  |  |  |  |  |  |  |  |  |  |  |  |  |  |  |  |  |  |  |  |
| 6. Lack of Spontaneity |  | Pearson's r |  | 0.605 | ** | 0.635 | ** | 0.721 | *** | 0.595 | ** | 0.466 | * | — |  |  |  |  |  |  |  |  |  |  |  |  |  |  |  |  |  |  |  |
|  |  | p-value |  | 0.005 |  | 0.003 |  | < .001 |  | 0.006 |  | 0.038 |  | — |  |  |  |  |  |  |  |  |  |  |  |  |  |  |  |  |  |  |  |
| 7. Stereotyped Thinking |  | Pearson's r |  | 0.524 | * | 0.660 | ** | 0.714 | *** | 0.818 | *** | 0.570 | ** | 0.717 | *** | — |  |  |  |  |  |  |  |  |  |  |  |  |  |  |  |  |  |
|  |  | p-value |  | 0.018 |  | 0.002 |  | < .001 |  | < .001 |  | 0.009 |  | < .001 |  | — |  |  |  |  |  |  |  |  |  |  |  |  |  |  |  |  |  |
| 8. Motor Retardation |  | Pearson's r |  | 0.714 | *** | 0.663 | ** | 0.690 | *** | 0.702 | *** | 0.346 |  | 0.720 | *** | 0.601 | ** | — |  |  |  |  |  |  |  |  |  |  |  |  |  |  |  |
|  |  | p-value |  | < .001 |  | 0.001 |  | < .001 |  | < .001 |  | 0.136 |  | < .001 |  | 0.005 |  | — |  |  |  |  |  |  |  |  |  |  |  |  |  |  |  |
| 9. Social Avoidance |  | Pearson's r |  | 0.385 |  | 0.580 | ** | 0.233 |  | 0.804 | *** | 0.133 |  | 0.182 |  | 0.528 | * | 0.451 | * | — |  |  |  |  |  |  |  |  |  |  |  |  |  |
|  |  | p-value |  | 0.093 |  | 0.007 |  | 0.322 |  | < .001 |  | 0.575 |  | 0.443 |  | 0.017 |  | 0.046 |  | — |  |  |  |  |  |  |  |  |  |  |  |  |  |
| 10. Facial expressivity |  | Pearson's r |  | -0.045 |  | 0.183 |  | 0.053 |  | -0.101 |  | 0.328 |  | 0.108 |  | 0.137 |  | -0.095 |  | -0.102 |  | — |  |  |  |  |  |  |  |  |  |  |  |
|  |  | p-value |  | 0.854 |  | 0.452 |  | 0.828 |  | 0.681 |  | 0.171 |  | 0.661 |  | 0.575 |  | 0.697 |  | 0.678 |  | — |  |  |  |  |  |  |  |  |  |  |  |
| 11. Vocal intensity |  | Pearson's r |  | -0.302 |  | -0.530 | * | -0.337 |  | -0.512 | * | -0.366 |  | -0.416 |  | -0.279 |  | -0.540 | * | -0.425 |  | 0.364 |  | — |  |  |  |  |  |  |  |  |  |
|  |  | p-value |  | 0.208 |  | 0.019 |  | 0.159 |  | 0.025 |  | 0.123 |  | 0.077 |  | 0.248 |  | 0.017 |  | 0.070 |  | 0.126 |  | — |  |  |  |  |  |  |  |  |  |
| 12. Fundamental frequency |  | Pearson's r |  | -0.355 |  | -0.632 | ** | -0.480 | * | -0.514 | * | -0.444 |  | -0.509 | * | -0.346 |  | -0.564 | * | -0.453 |  | 0.184 |  | 0.935 | *** | — |  |  |  |  |  |  |  |
|  |  | p-value |  | 0.136 |  | 0.004 |  | 0.037 |  | 0.024 |  | 0.057 |  | 0.026 |  | 0.147 |  | 0.012 |  | 0.051 |  | 0.451 |  | < .001 |  | — |  |  |  |  |  |  |  |
| 13. Fundamental frequency |  | Pearson's r |  | -0.192 |  | -0.447 |  | -0.071 |  | -0.388 |  | -0.030 |  | -0.280 |  | -0.170 |  | -0.365 |  | -0.404 |  | 0.179 |  | 0.581 | ** | 0.529 | * | — |  |  |  |  |  |
|  |  | p-value |  | 0.432 |  | 0.055 |  | 0.774 |  | 0.101 |  | 0.904 |  | 0.246 |  | 0.485 |  | 0.124 |  | 0.086 |  | 0.464 |  | 0.009 |  | 0.020 |  | — |  |  |  |  |  |
| 14. Harmonics to noise ratio |  | Pearson's r |  | -0.228 |  | -0.668 | ** | -0.583 | ** | -0.495 | * | -0.413 |  | -0.480 | * | -0.385 |  | -0.481 | * | -0.303 |  | 0.174 |  | 0.654 | ** | 0.774 | *** | 0.476 | * | — |  |  |  |
|  |  | p-value |  | 0.347 |  | 0.002 |  | 0.009 |  | 0.031 |  | 0.079 |  | 0.038 |  | 0.104 |  | 0.037 |  | 0.207 |  | 0.475 |  | 0.002 |  | < .001 |  | 0.039 |  | — |  |  |  |
| 15. Vocal jitter |  | Pearson's r |  | 0.057 |  | 0.564 | * | 0.457 | * | 0.353 |  | 0.333 |  | 0.328 |  | 0.209 |  | 0.308 |  | 0.209 |  | -0.097 |  | -0.541 | * | -0.691 | ** | -0.278 |  | -0.937 | *** | — |  |
|  |  | p-value |  | 0.816 |  | 0.012 |  | 0.049 |  | 0.138 |  | 0.164 |  | 0.170 |  | 0.391 |  | 0.200 |  | 0.390 |  | 0.692 |  | 0.017 |  | 0.001 |  | 0.249 |  | < .001 |  | — |  |
| 16. Speech to silence ratio |  | Pearson's r |  | -0.396 |  | -0.510 | * | -0.468 | * | -0.442 |  | -0.453 |  | -0.541 | * | -0.326 |  | -0.552 | * | -0.358 |  | 0.161 |  | 0.869 | *** | 0.923 | *** | 0.260 |  | 0.575 | ** | -0.510 | * |
|  |  | p-value |  | 0.093 |  | 0.026 |  | 0.043 |  | 0.058 |  | 0.051 |  | 0.017 |  | 0.174 |  | 0.014 |  | 0.133 |  | 0.510 |  | < .001 |  | < .001 |  | 0.283 |  | 0.010 |  | 0.026 |  |
